# Transcriptomic profiling of cerebrospinal fluid predicts shunt surgery responses in patients with Normal Pressure Hydrocephalus

**DOI:** 10.1101/2022.08.20.22279018

**Authors:** Zachary Levin, Owen P. Leary, Victor Mora, Shawn Kant, Sarah Brown, Konstantina Svokos, Umer Akbar, Thomas Serre, Petra Klinge, Alexander Fleischmann, Maria Grazia Ruocco

**Author notes:** Correspondence should be addressed to M.G.R. or A.F.

## Abstract

Molecular biomarkers for neurodegenerative diseases are critical for advancing diagnosis and therapy. Normal Pressure Hydrocephalus (NPH) is a neurodegenerative disorder characterized by progressive gait impairment, urinary incontinence, and cognitive decline. In contrast to most other neurodegenerative disorders, NPH symptoms can be improved by the placement of a ventricular shunt that drains excess cerebrospinal fluid (CSF). A major challenge in NPH management is the identification of patients who benefit from shunt surgery. Here, we perform genome-wide RNA sequencing of extracellular vesicles in CSF of 42 NPH patients, and we identify genes and pathways whose expression levels correlate with gait, urinary or cognitive symptom improvement after shunt surgery. We describe a machine learning algorithm trained on these gene expression profiles that can predict shunt surgery response with high accuracy. The transcriptomic signatures we identified have important implications for improving NPH diagnosis and treatment and for understanding disease etiology.

## Introduction

Neurodegenerative diseases represent a large group of neurological disorders caused by the loss of neurons, typically resulting in progressive cognitive, motor, and autonomic dysfunction^1, 2^. Despite major efforts to address the growing health crisis caused by neurodegenerative diseases, reliable diagnostic tests and effective therapies remain largely unavailable^3, 4^.

Normal Pressure Hydrocephalus (NPH) is a progressive neurodegenerative disorder affecting an estimated 20 million people worldwide^5–7^. It is characterized by the accumulation of cerebrospinal fluid (CSF) in the brain ventricles and results in progressive locomotor dysfunction, urinary incontinence, and cognitive decline^8, 9^. NPH has been estimated to account for 5-10% of all dementia cases^6, 7,10^. However, because symptoms overlap with those of other neurodegenerative disorders including Alzheimer’s and Parkinson’s Disease, accurately differentiating NPH from other neurodegenerative disorders remains challenging^8, 11^.

In contrast to other neurodegenerative disorders, NPH patients can be treated by shunt surgery, the placement of a ventriculo-peritoneal shunt to drain excess CSF. Shunt surgery can dramatically ameliorate and even reverse patients’ symptoms and prevent progression into dementia^8, 12^. However, diagnostic tools to accurately predict responses to shunt surgery are limited. Diagnostic tests have been developed to simulate the response to CSF drainage by utilizing a high-volume CSF tap tests or a lumbar CSF drainage. While the positive predictive value of those tests can reach up to 80% in specialized NPH clinics, their negative predictive value has remained critically low^13–15^. Furthermore, shunt surgery is associated with postoperative risks including shunt infection and should be limited to patients who experience symptom improvement^14, 16^. Therefore, there is a critical need for an objective, quantitative diagnostic test that can accurately identify patients who most likely benefit from shunt surgery.

Cerebrospinal fluid (CSF) is an attractive source for the identification of new NPH biomarkers. CSF surrounds the brain and spinal canal and performs vital homeostatic functions including nourishment, waste removal, and mechanical protection^17–19^. CSF contains a large variety of metabolites and extracellular signaling molecules, including growth factors, hormones, lipids, and RNAs. Importantly, pathological states including neurodegeneration lead to alterations in the molecular composition of CSF^20, 21^. Several recent studies suggested that core biomarkers for neurodegerenation and inflammation, such as β-amyloid and tau proteins, IL-6, IL-8, or TNFα have limited predictive value for shunt surgery outcomes^22–26^.

Therefore, to identify new CSF-based biomarkers for NPH diagnosis and therapy, we collected CSF from 42 NPH patients during placement of a ventriculoperitoneal shunt. We analyzed the expression levels of core molecular markers for neurodegeneration, and we performed transcriptomic profiling of extracellular vesicles in CSF. We identified genes and pathways whose expression levels correlate with the presence of urinary and cognitive symptoms before shunt surgery, and with gait, urinary and cognitive symptom improvement after shunt surgery. We then developed a machine learning pipeline to identify genes that are highly predictive of shunt surgery outcomes. Together, our data provide evidence for the presence of RNA biomarkers in the CSF of NPH patients that can predict shunt surgery benefits with high accuracy.

## Results

### Demographics of patient cohort and study design

We collected ventricular CSF samples from 42 NPH patients who underwent shunt surgery at the Center for CSF Disorders of the Brain and Spine at Rhode Island Hospital (**Table 1**). The median age of the patient cohort was 73 (range: 60-89) and 19 (45%) of the patients were male. Thirty-nine of the patients were diagnosed with idiopathic NPH (iNPH) and 3 had secondary NPH (sNPH). Two of the sNPH patients developed NPH following traumatic brain injury. The third sNPH patient was diagnosed with NPH 20 years after resection of a low-grade astrocytoma with subsequent chemoradiation, and after serial imaging demonstrated disproportionate ventricular enlargement without evidence of residual or recurrent tumor over that period. All patients (42/42) showed ventriculomegaly on magnetic resonance imaging (MRI) and reported gait impairment. Thirty-five of 42 patients (83%) reported urinary incontinence, and 36/42 patients (86%) had cognitive impairment (see **supplementary table 1** for combinations of symptoms). The mean baseline modified Rankin Scale (mRS) score of the cohort was 2.50 (± 0.83). In 12 patients, a high-volume (50 mL) lumbar tap test was performed to support the candidacy for shunt placement by an observed improvement in walking speed and stride length post lumbar tap. Thirty-eight of the 42 patients (90%) reported to the 3-month postoperative follow-up appointment. Four patients (10%) showed improvement in all symptoms, 22 (58%) reported improvement in balance/gait symptoms, 19 (61%) reported improvement in urinary incontinence, and 20 patients (62%) reported improvement in cognitive impairment. The mean mRS score at 3-months post shunting was 2.24 (± 0.88) (**Table 1** and **supplementary table 1**).

**Table 1.**
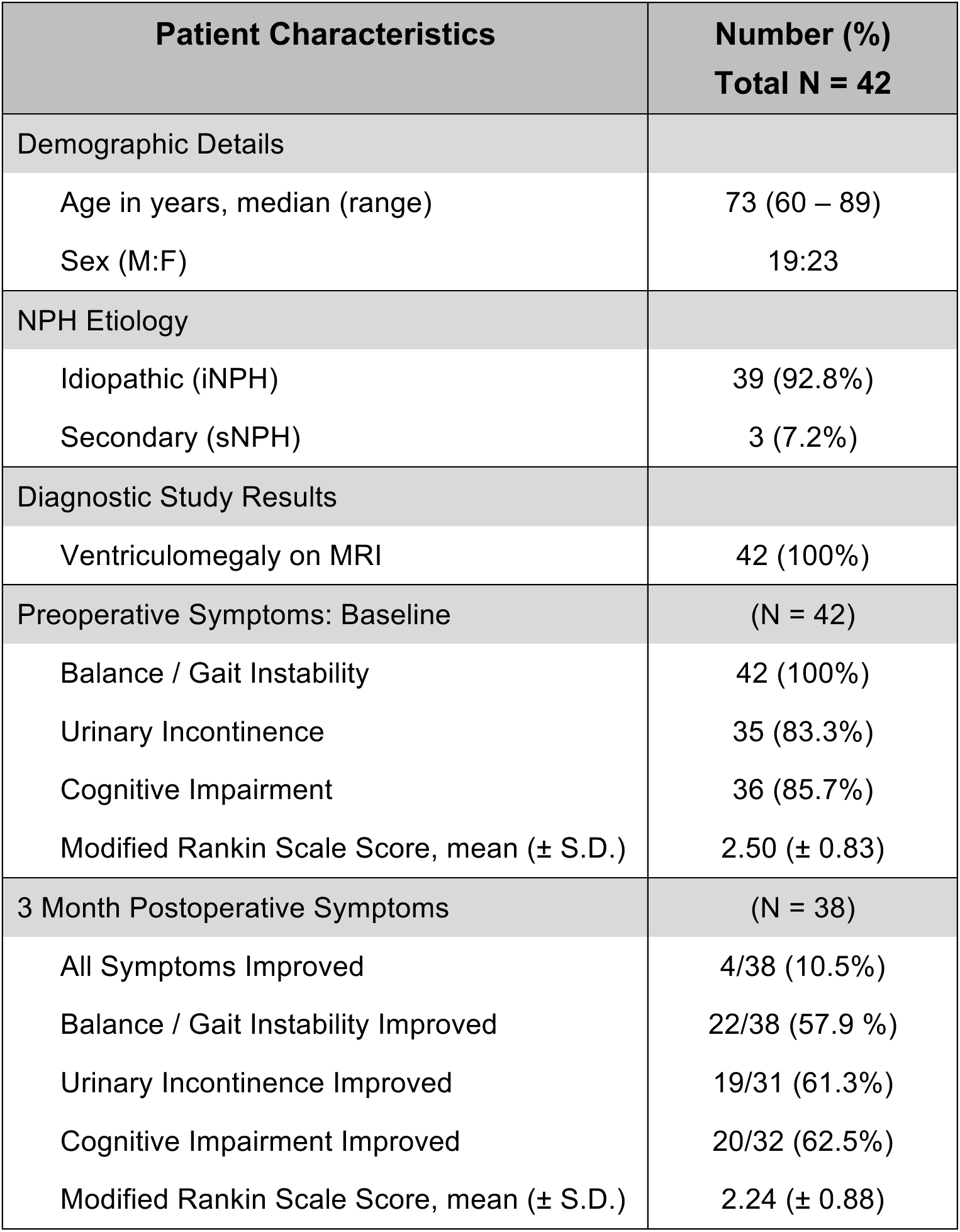
Patient demographics, baseline NPH symptoms, and 3-month shunt surgery responses. 42 CSF samples were collected and 38 patients returned 3-month after shunt surgery for follow-up appointments. For calculating the percent of patients that improved in each symptom category after surgery, the number of patients reporting symptom improvement was divided by the number of patients that initially presented with that symptom.

| **Patient Characteristics** | **Number (%)**<br>**Total N = 42** |
| --- | --- |
| **Demographic Details** |  |
| Age in years, median (range) | 73 (60 – 89) |
| Sex (M:F) | 19:23 |
| **NPH Etiology** |  |
| Idiopathic (iNPH) | 39 (92.8%) |
| Secondary (sNPH) | 3 (7.2%) |
| **Diagnostic Study Results** |  |
| Ventriculomegaly on MRI | 42 (100%) |
| **Preoperative Symptoms: Baseline** | **(N = 42)** |
| Balance / Gait Instability | 42 (100%) |
| Urinary Incontinence | 35 (83.3%) |
| Cognitive Impairment | 36 (85.7%) |
| Modified Rankin Scale Score, mean (± S.D.) | 2.50 (± 0.83) |
| **3 Month Postoperative Symptoms** | **(N = 38)** |
| All Symptoms Improved | 4/38 (10.5%) |
| Balance / Gait Instability Improved | 22/38 (57.9 %) |
| Urinary Incontinence Improved | 19/31 (61.3%) |
| Cognitive Impairment Improved | 20/32 (62.5%) |
| Modified Rankin Scale Score, mean (± S.D.) | 2.24 (± 0.88) |

CSF samples were split into aliquots used for ELISA immunoassay and transcriptome analysis. For transcriptome analysis, extracellular vesicles were isolated and mRNA extracted before Illumina RNA sequencing. RNA sequencing data was analyzed to identify differentially expressed genes and pathways, and a Support Vector Machine classifier was used to predict 3-months shunt surgery responses based on pre- and postoperative clinical assessments (**Figure 1**).

**Figure 1.**
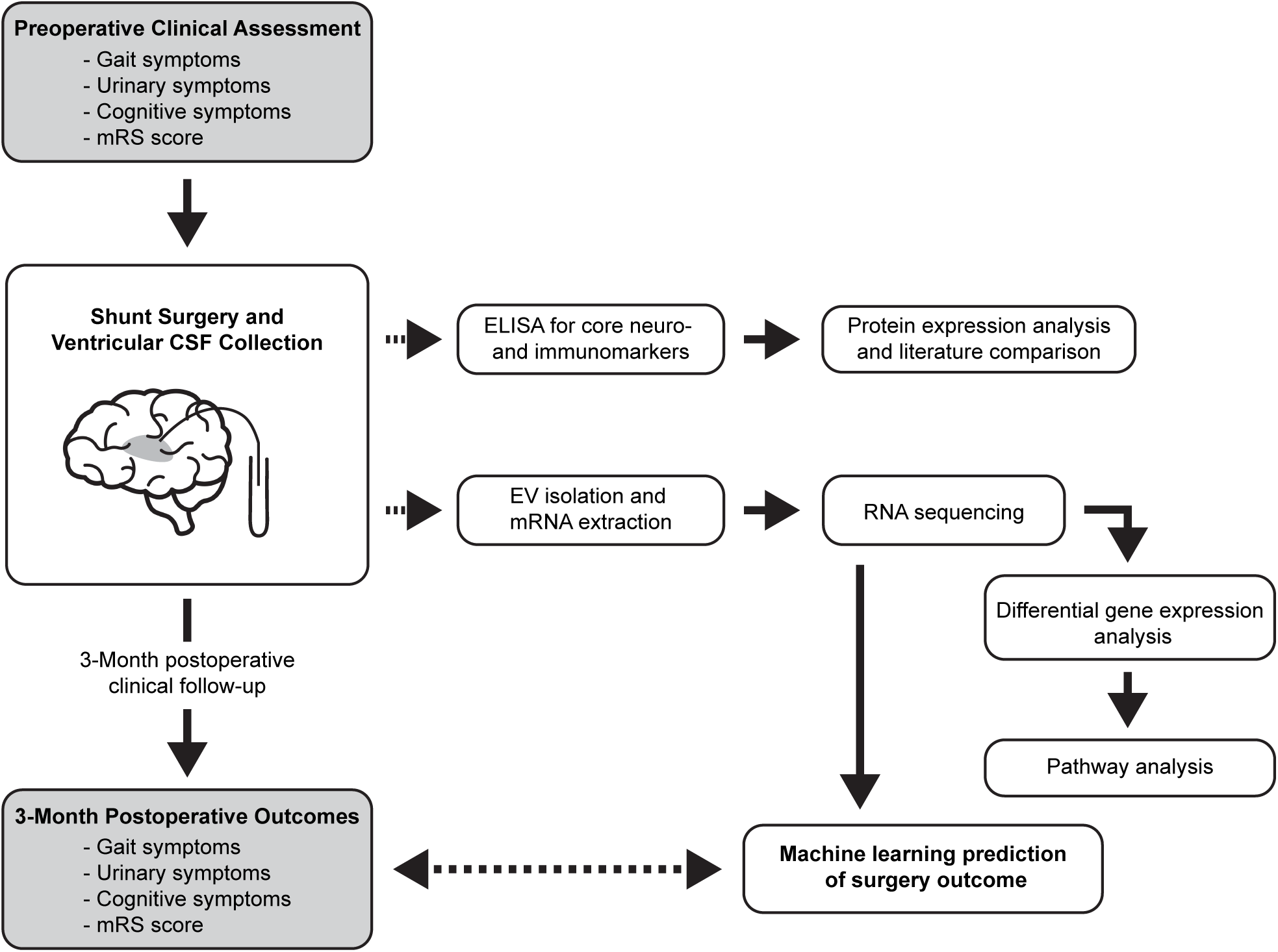
Study design.

### Expression levels of core CSF biomarkers for neurodegeneration do not correlate with NPH shunt surgery responses

Many neurodegenerative disorders including NPH are characterized by alterations in the CSF expression levels of β-amyloid and tau proteins, and increased levels of neuroinflammatory markers including IL-6, IL-8, and TNFα^23, 24, 27–31^. Furthermore, increased expression of markers for neuroaxonal damage (neurofilament light chain, NfL) and glial activation (glial fibrillary acidic protein, GFAP) are common features of neurodegenerative disorders^22, 32–35^.

We therefore tested whether the CSF expression levels of these core CSF biomarkers for neurodegeneration correlated with gait, urinary or cognitive symptom improvement after shunt surgery in our patient cohort. We found that β-amyloid 42 (Aβ42) and p-tau181 expression, and the p-tau181/total-tau ratio, were similar in patients who showed symptom improvement compared to patients who did not show symptom improvement (two-sided Mann-Whitney U-test with Benjamini-Hochberg correction, all p > 0.05, **Figure 2a-c**). Similarly, IL-6, IL-8 and TNFα levels were similar across patient groups (two-sided Mann-Whitney U-test with Benjamini-Hochberg correction, all p > 0.05, **Figure 2d-f**). Finally, NfL and GFAP expression levels were similar across patient groups (two-sided Mann-Whitney U-test with Benjamini-Hochberg correction, all p > 0.05, **Figure 2g, h**). Thus, the expression levels of the core neurodegeneration markers Aβ42, p-tau181, IL-6, IL-8, TNFα, NfL and GFAP did not correlate with shunt surgery responses in our cohort.

**Figure 2.**
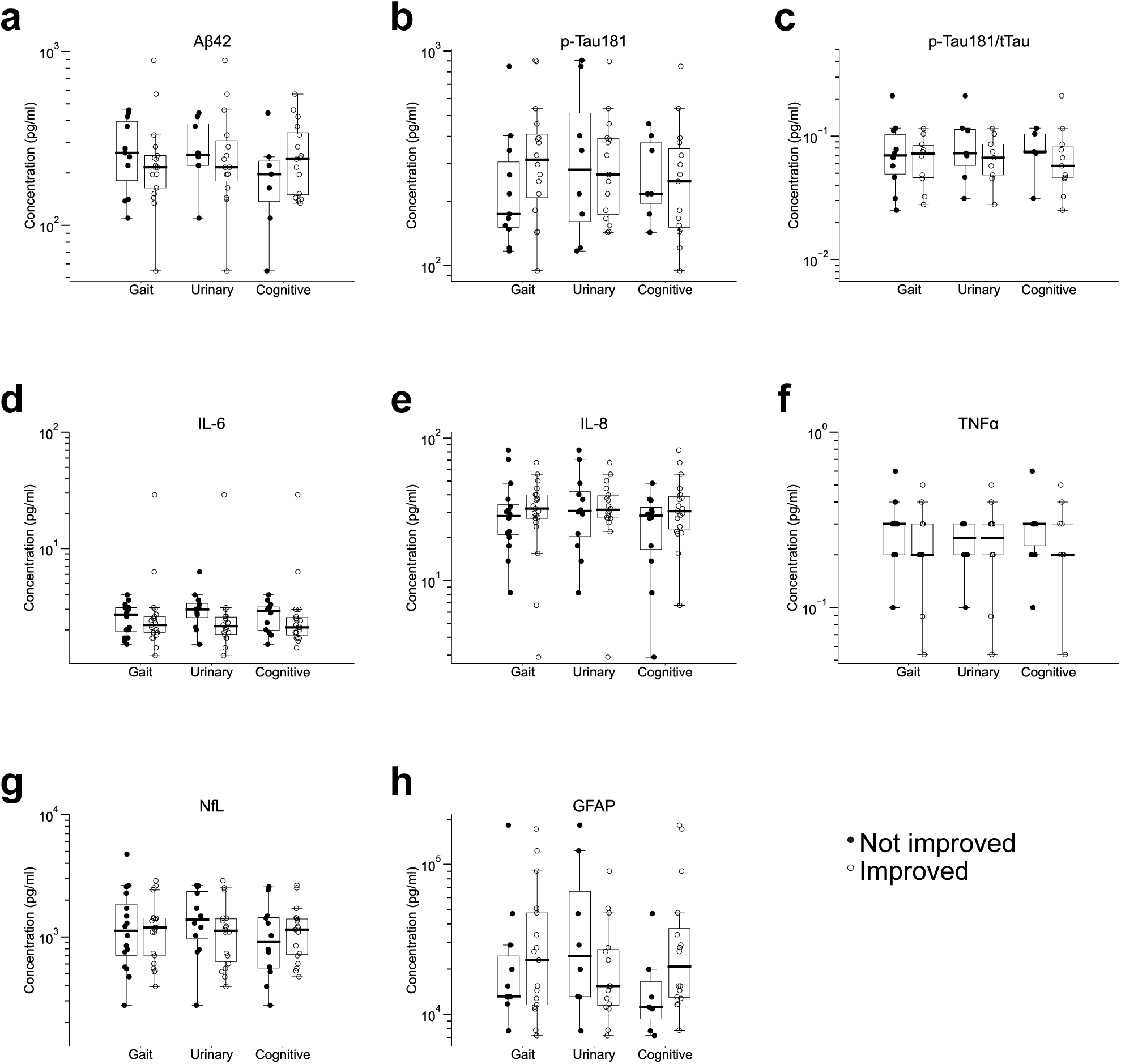
Concentrations of core CSF biomarkers for neurodegeneration do not correlate with NPH shunt surgery responses. **a.** Concentrations of Aβ42 for patients with improvement (open circles) or without improvement (filled circles) in gait, urinary, and cognitive symptoms after shunting. All U- and p-values calculated by two-sided Mann-Whitney U-test with Benjamini-Hochberg multiple hypothesis correction (Gait, U = 74.5, p = 0.35; Urinary, U = 48, p = 0.37; Cognitive, U = 41, p = 0.35). **b.** Concentrations of p-Tau181 for patients with or without improvement in gait, urinary, and cognitive symptoms after shunting. (Gait, U = 59.5, p = 0.35; Urinary, U = 59.5, p = 0.5; Cognitive, U = 46, p = 0.45). **c.** Ratio of concentrations of p-Tau181 over total tau (tTau) for patients with or without improvement in gait, urinary, and cognitive symptoms after shunting. (Gait, U = 47, p = 0.49; Urinary, U = 24, p = 0.37; Cognitive, U = 21, p = 0.37). **d.** Concentrations of IL-6 for patients with or without improvement in gait, urinary, and cognitive symptoms after shunting. (Gait, U = 138, p = 0.35; Urinary, U = 52.5, p = 0.24; Cognitive, 81.5, p = 0.35). **e.** Concentrations of IL-8 for patients with or without improvement in gait, urinary, and cognitive symptoms after shunting. (Gait, U = 137, p = 0.35; Urinary, U = 105, p = 0.49; Cognitive, U = 85, p = 0.35). **f.** Concentrations of TNFα for patients with or without improvement in gait, urinary, and cognitive symptoms after shunting. (Gait, U = 50.5, p = 0.35; Urinary, U = 48.5, p = 0.49; Cognitive, U = 42, p = 0.35). **g.** Concentrations of NfL for patients with or without improvement in gait, urinary, and cognitive symptoms after shunting. (Gait, U = 165, p = 0.49; Urinary, U = 76, p = 0.35; Cognitive, U = 100.5, p = 0.42). **h.** Concentrations of GFAP for patients with or without improvement in gait, urinary, and cognitive symptoms after shunting. (Gait, U = 85, p = 0.45; Urinary, U = 44, p = 0.35; Cognitive, U = 26, p = 0.29).

### CSF transcriptome analysis identifies molecular signatures of urinary incontinence and cognitive impairment

NPH patients typically report different combinations of gait, urinary, and cognitive symptoms ^8, 9, 36^. We therefore examined whether the patients’ CSF contained transcriptomic signatures that correlated with preoperative NPH symptoms. In this NPH cohort, all patients reported gait impairment. We thus examined whether extracellular vesicles in the CSF contained signatures for urinary incontinence (83% of patients) and cognitive impairment (86% of patients).

Differential gene expression analysis identified 52 genes whose expression was significantly upregulated in patients with urinary incontinence (all Benjamini-Hochberg adjusted p < 0.05, **Figure 3a**). To visualize gene expression on a per patient basis, we plotted the normalized expression of the top differentially expressed gene by patient. We found that expression of *MOK* was significantly increased in patients with urinary symptoms compared to patients without urinary symptoms (two-sided Mann-Whitney U-test, p = 0.011, **Figure 3b)**. By grouping genes into biologically meaningful gene sets, pathway analysis can reduce noise in transcriptome data and allow for a more robust characterization of molecular changes ^37^. We therefore performed pathway analysis using the Kyoto Encyclopedia of Genes and Genomes (KEGG) pathways, and we identified 12 metabolic pathways that were significantly enriched in patients with urinary incontinence (FDR < 0.25, **Figure 3c**).

**Figure 3.**
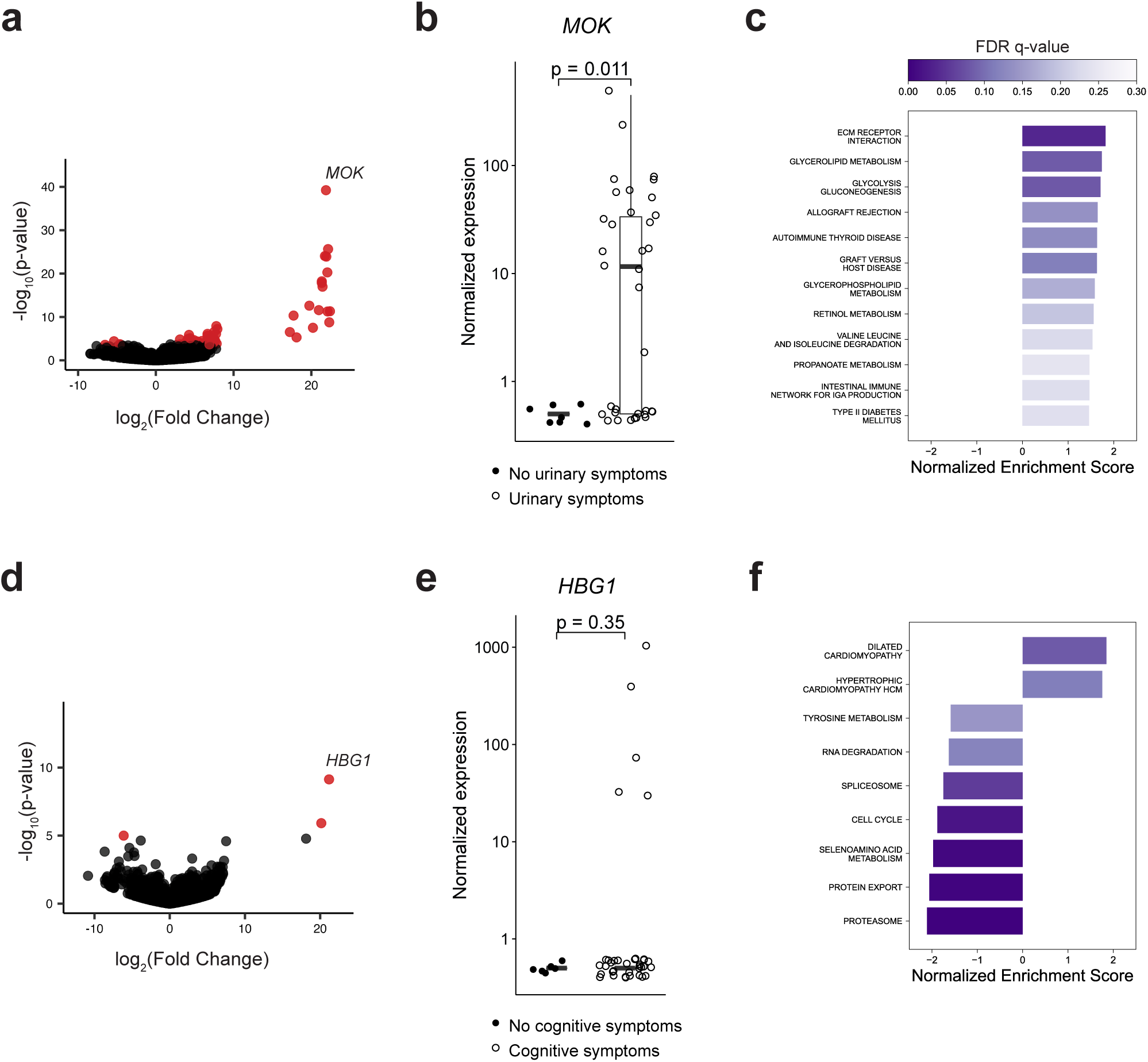
CSF signatures for urinary incontinence and cognitive impairment in NPH patients. **a.** Differentially expressed genes (DEGs) correlated with urinary incontinence. Positive fold change indicates increased expression in patients with urinary incontinence. Red dots indicate genes with Benjamini-Hochberg adjusted p-value < 0.05. **b.** Normalized expression of *MOK* for each patient. Filled circles: patients without urinary incontinence (n = 7), open circles: patients with urinary incontinence (n = 35). U- and p-values calculated by two-sided Mann-Whitney U-test (U = 52.5, p = 0.011). **c.** KEGG pathways enriched in patients with urinary incontinence. Positive normalized enrichment scores (NES) indicate upregulation in patients with urinary incontinence. Color of each bar indicates false discovery rate (FDR) q-value. **d.** Plot as in (a) showing DEGs based on cognitive symptoms in NPH patients. Red dots indicate genes with Benjamini-Hochberg adjusted p-value < 0.05. **e.** Normalized expression of *HBG1* for each patient. Filled circles: patients without cognitive impairment (n = 6), open circles: patients with cognitive impairment (n = 36). U- and p-values calculated by two-sided Mann-Whitney U-test (U = 93, p = 0.35). **f.** KEGG pathways upregulated (positive NES) and downregulated (negative NES) in patients with cognitive symptoms. Color of each bar indicates FDR q-value.

Differential gene expression analysis identified 3 significantly dysregulated genes in patients with cognitive impairment (all Benjamini-Hochberg adjusted p < 0.05, **Figure 3d**). However, on a per patient basis, the expression of *HBG1*, the top differentially expressed gene, was not statistically different between patients with and without cognitive symptoms (two-sided Mann-Whitney U-test, p = 0.35, **Figure 3e**). Finally, KEGG pathway analysis identified 9 metabolic pathways that were significantly dysregulated in patients with cognitive impairment (FDR < 0.25, **Figure 3f**). Together, these data identify genes and pathways whose CSF expression levels correlate with the presence of urinary and cognitive symptoms in NPH patients.

### CSF transcriptome analysis identifies molecular signatures of NPH shunt surgery responses

We next tested whether CSF gene expression profiles correlated with NPH shunt surgery responses. We performed differential gene expression and pathway analysis for patients who did or did not show symptom improvement 3 months after shunt surgery. We examined three independent, binary outcomes: gait improvement, urinary improvement, and cognitive improvement (**Figure 4**).

**Figure 4.**
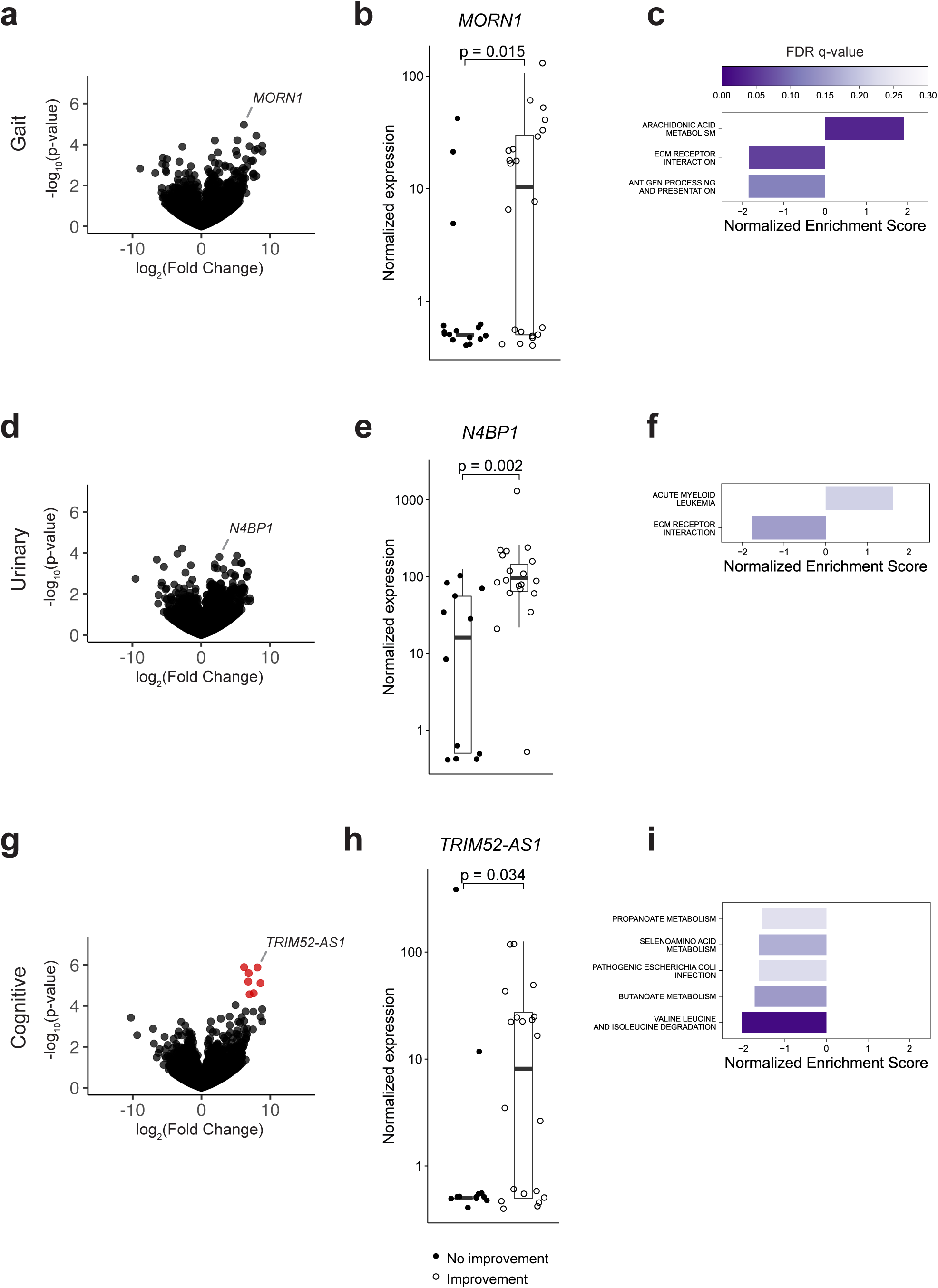
CSF signatures for gait, urinary, and cognitive symptom improvement after shunt surgery. **a.** Differentially expressed genes (DEGs) correlated with gait symptom improvement 3 months after shunt surgery. Positive fold change indicates increased expression in patients with gait improvement. **b.** Normalized expression of *MORN1* for each patient. Filled circles: patients without gait improvement (n = 16), open circles: patients with gait improvement (n = 22). U- and p-values calculated by two-sided Mann-Whitney U-test (U = 101.5, p = 0.015). **c.** KEGG pathways enriched in patients with gait improvement. Positive normalized enrichment scores (NES) indicate upregulation in patients with gait improvement. Color of each bar indicates FDR q-value. **d.** DEGs correlated with urinary symptom improvement 3 months after shunt surgery. Positive fold change indicates increased expression in patients with urinary symptom improvement. **e.** Normalized expression of *N4BP1* for each patient. Filled circles: patients without urinary symptom improvement (n = 12), open circles: patients with urinary symptom improvement (n = 19). U- and p-values calculated by two-sided Mann-Whitney U-test (U = 37.5, p = 0.002). **f.** KEGG pathways enriched in patients with urinary symptom improvement. Color of each bar indicates FDR q-value. **g.** DEGs correlated with cognitive symptom improvement 3 months after shunt surgery. Positive fold change indicates increased expression in patients with cognitive improvement. Red dots indicate genes with Benjamini-Hochberg adjusted p-value < 0.05. **h.** Normalized expression of *TRIM52-AS1* for each patient. Filled circles: patients without cognitive improvement (n = 12), open circles: patients with cognitive improvement (n = 20). U- and p-values calculated by two-sided Mann-Whitney U-test (U = 70, p = 0.034). **i.** KEGG pathways enriched in patients with cognitive improvement. Color of each bar indicates FDR q-value.

For patients with improvement in gait symptoms, differential gene expression analysis did not identify individual genes that reached statistical significance after multiple hypothesis correction (Benjamini-Hochberg adjusted p < 0.05, **Figure 4a**). Despite the lack of statistically significant differentially expressed genes, we plotted the normalized expression of the gene with lowest adjusted p-value for each patient to visualize gene expression on a per patient basis. We found that patients with gait improvement had increased expression of *MORN1* (two-sided Mann-Whitney U-test, p = 0.015, **Figure 4b**). Furthermore, KEGG pathway analysis identified 3 pathways that were significantly dysregulated in patients with gait improvement (FDR < 0.25, **Figure 4c**).

For patients with improvement in urinary incontinence, differential gene expression analysis did not identify individual genes that reached statistical significance (Benjamini-Hochberg adjusted p < 0.05 **Figure 4d**). Nevertheless, expression of *N4BP1* was higher in patients with urinary improvement (two-sided Mann-Whitney U-test, p = 0.002, **Figure 4e**). KEGG pathway analysis identified 2 pathways that were significantly dysregulated in patients with urinary improvement (FDR < 0.25, **Figure 4f**).

Finally, for patients with cognitive improvement after shunt surgery, differential gene expression analysis identified 7 significantly upregulated genes (all Benjamini-Hochberg adjusted p < 0.05, **Figure 4g** and **supplementary table 2**), and patients with cognitive improvement showed significantly increased expression of *TRIM52-AS1* (two-sided Mann-Whitney U-test, p = 0.034, **Figure 4h**). KEGG pathway analysis identified 5 metabolic pathways that were significantly downregulated in patients with cognitive improvement (FDR < 0.25, **Figure 4i**). Together, these data indicate widespread changes, with broadly varying effect sizes, in the expression of genes and pathways that correlate with improved gait, urinary, and cognitive symptoms after NPH shunt surgery.

### A machine learning classifier accurately predicts shunt surgery responses based on CSF transcriptomes

To test if the changes in CSF gene and pathway expression we identified can predict shunt surgery responses, we developed a machine learning pipeline. Using a leave-one-out cross-validation procedure, we first z-scored gene expression and fitted a logistic regression model to the gene expression data to identify predictive genes as those with the highest weights assigned by the model. Next, we trained a Support Vector Machine (SVM) classifier to predict symptom improvement after shunt surgery based on these genes. We then determined the optimal number of genes to predict symptom improvement by varying the number of genes selected in each classifier fold between 10 and 1000. Finally, we calculated the area under the receiver operating characteristic curver (AUC) of the SVM classifier trained on the most predictive genes (**Figure 5**). For gait improvement, classifier performance peaked when 30 genes were selected, and a SVM classifier fitted to the expression of the selected genes predicted gait improvement with an AUC = 0.80 (**Figure 6a, b**). To visualize class separation, we performed PCA on the genes selected in the first iteration of the classifier, which revealed clear separation of patient groups by clinical outcome (**Figure 6c**). To predict urinary improvement, classifier performance peaked with 140 genes and an AUC = 0.79 (**Figure 6d** and **e**). Plotting the samples along the first two principal components in PCA space revealed clear separation of patient groups by clinical outcome (**Figure 6f**). Finally, we found that 50 genes selected from the logistic regression model best predicted cognitive improvement (**Figure 6g**). A SVM classifier fitted to the expression of 50 genes predicted cognitive improvement with an AUC = 0.81 (**Figure 6h**), consistent with clear separation of patient groups in PCA space (**Figure 6I**). Together, these results suggest that the CSF of NPH patients in our cohort contains transcriptomic profiles that robustly distinguish patients based on their response to shunt surgery. Furthermore, a small (∼100) set of genes is sufficient to predict gait, urinary, and cognitive symptom improvement.

**Figure 5.**
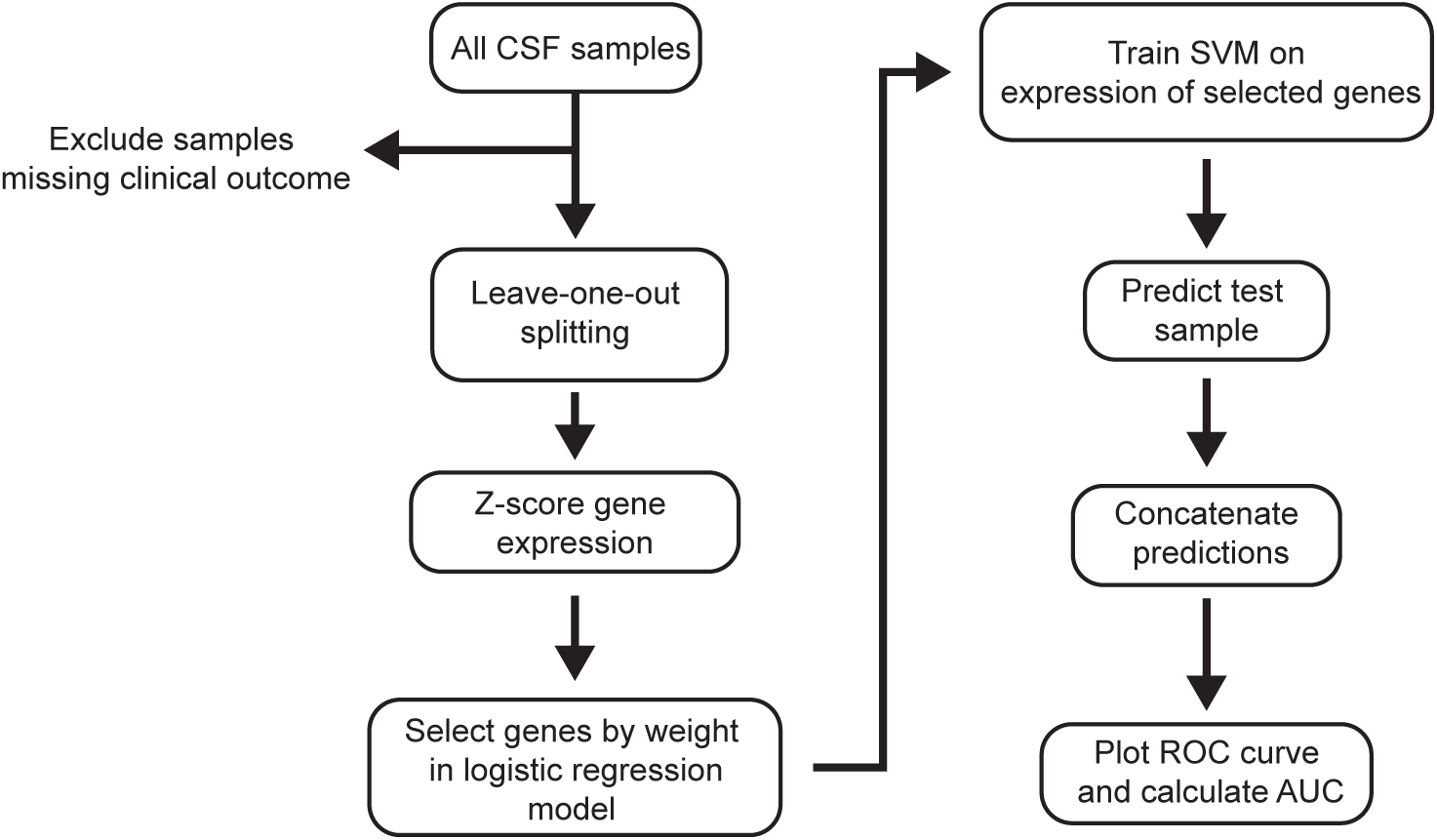
Feature selection by logistic regression weights and SVM machine learning approach for predicting symptom improvement after shunt surgery.

**Figure 6.**
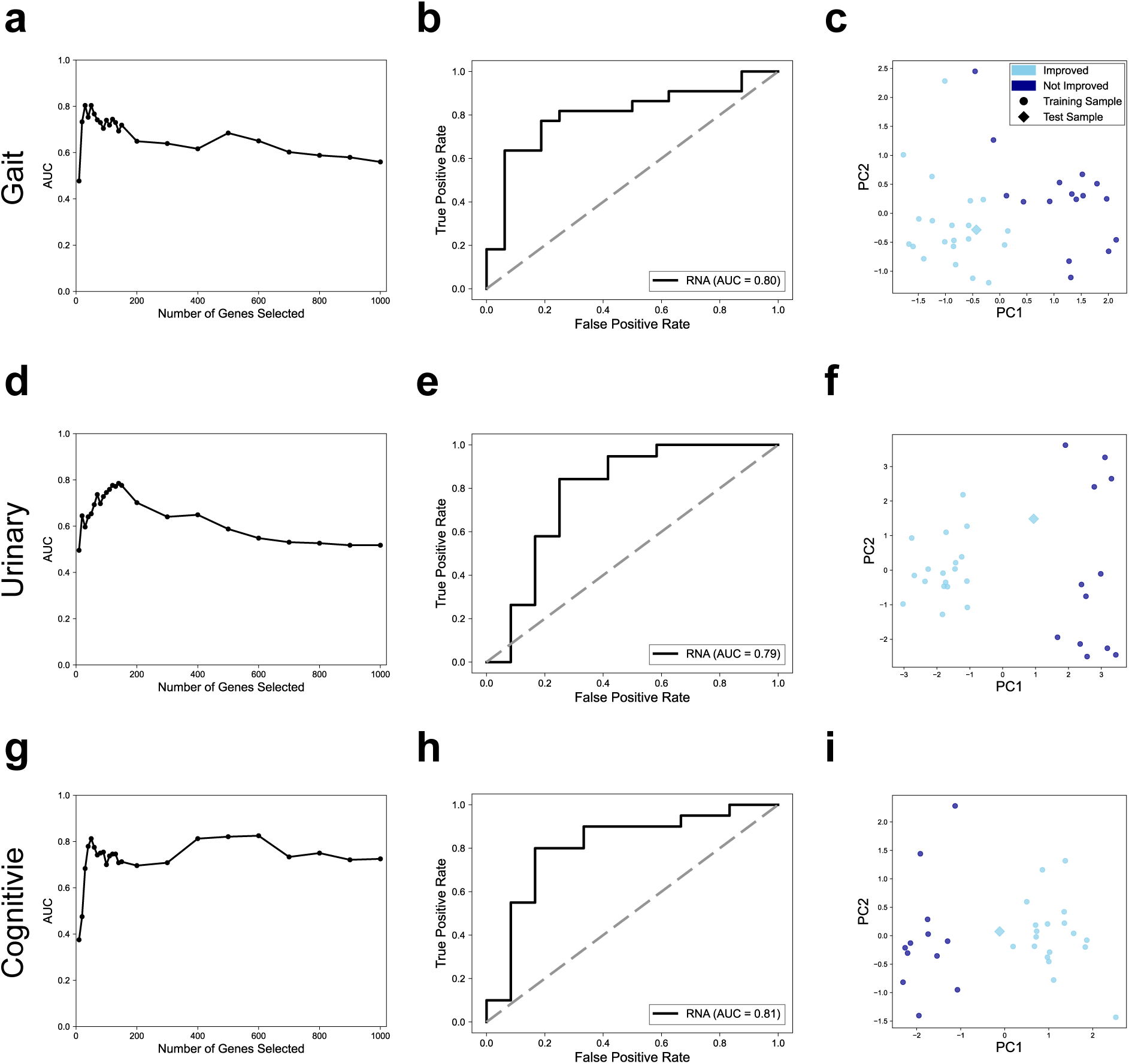
A machine learning classifier trained on CSF transcriptomic profiles accurately predicts symptom improvement after shunt surgery. **a.** Effect of number of genes selected in each classifier fold on predicting gait improvement. **b.** ROC curve for predicting gait improvement using 30 genes selected by weights in a logistic regression model. Solid line indicates prediction based on selected genes, broken line: chance level. **c.** PCA plot fit to the training set expression of genes selected when the first sample is the test set. Circles: training set samples. Diamond: test set sample. Light blue: patients with gait improvement. Dark blue: patients without gait improvement. **d.** Effect of number of genes selected in each classifier fold on predicting urinary improvement. **e.** ROC curve for predicting urinary improvement using 140 genes selected by weights in a logistic regression model. Solid line indicates prediction based on selected genes. **f.** PCA plot fit to the training set expression of genes selected when the first sample is the test set. Circles: training set samples. Diamond: test set sample. Light blue: patients with urinary improvement. Dark blue: patients without urinary improvement. **g.** Effect of number of genes selected in each classifier fold on predicting cognitive improvement. **h.** ROC curve for predicting cognitive improvement using 50 genes selected by weights in a logistic regression model. Solid line indicates prediction based on selected genes. **i.** PCA plot fit to the training set expression of genes selected when the first sample is the test set. Circles: training set samples. Diamond: test set sample. Light blue: patients with cognitive improvement. Dark blue: patients without cognitive improvement.

## Discussion

Neurodegenerative diseases are a major, growing health and economic burden worldwide and there is an urgent need for improved diagnostic and therapeutic solutions^38^. Normal Pressure Hydrocephalus (NPH) is unique among dementia-causing disorders because it can be effectively treated by surgical shunt placement^8, 12, 13^. However, only a fraction of NPH patients who undergo shunt surgery experience symptom improvement, with estimates varying from 30% to 85%^2, 13–16^. In the present study, we report that the transcriptomes of extracellular vesicles (EVs) extracted from ventricular cerebrospinal fluid (CSF) contain molecular signatures that correlate with the clinical response to shunt surgery outcomes for the classical NPH symptoms (gait/balance impairment, urinary incontinence, and cognitive impairment). Using machine learning, we demonstrate that these molecular signatures support the accurate prediction of shunt surgery responses.

Our cohort of 42 patients represents some of the heterogeneity of clinical symptoms typically observed among NPH patients. Through differential gene expression and pathway analysis, we identified genes and pathways whose expression levels correlated with the presence of urinary and cognitive symptoms. These molecular signatures are also candidate biomarkers for the differential diagnosis of neurodegenerative disorders with clinical symptoms similar to NPH, including Alzheimer’s and Parkinson’s Disease^11, 23, 39^.

We also observed widespread changes in gene and pathway expression when comparing shunt surgery responders to non-responders. Based on these observations, we applied a machine learning approach to the CSF transcriptome data to predict shunt surgery responses based on CSF transcriptomic profiling. Our pipeline predicted improvements in gait, urinary, and cognitive symptoms with AUCs of ∼0.8, thus substantially improving upon earlier studies relying on common markers for neurodegeneration and inflammation^22–26^. Follow-up studies using CSF from lumbar puncture and with larger patient cohorts will be required to validate these findings, identify molecular signatures for combinations of symptoms, and to optimize gene sets for NPH diagnosis and shunt surgery prognosis. We envision that our findings will lead to the development of a simple and robust predictive test, which will improve clinical decision making and NPH patient care. Furthermore, being able to separately predict which domains of the NPH triad are likely to improve following shunting for a given patient would substantially improve the ability to weigh risk and benefit of surgery preoperatively in an individualized manner.

Our machine learning approach allowed us to identify candidate genes for shunt surgery responses and estimate the optimal number of genes required for accurate classification. Our results suggest that a small number of genes is sufficient for predicting NPH shunt surgery responses. Several of the top predictive genes selected by the classifier have previously been linked to neurodegeneration and play prominent roles in Wnt signaling, oxidative phosphorylation and proteasome function (**Supplementary Table 3**)^40–42^. Together with recently identified genetic risk factors for NPH^43, 44^, these genes and pathways provide critical new entry points for the investigation of molecular mechanisms underlying NPH etiology.

## Methods

### Patient cohort recruitment and clinical follow-up

Following recruitment of a pilot cohort of 3 patients in November 2019, a total of 54 patients with Normal Pressure Hydrocephalus (NPH) were approached in a cerebrospinal fluid (CSF) disorders specialty neurosurgical clinic at Rhode Island Hospital between 1/1/2020 and 6/1/2021 and invited to participate in the study. Six declined to participate and 48 agreed, providing informed consent either directly or with the aid of a legally authorized representative, as per Rhode Island Hospital Institutional Review Board (IRB) authorization (#1492994). CSF samples were collected from 42 patients. 38/42 patients returned for a 3 (+/-1) month clinical follow-up postoperatively (**Table 1**).

### Clinical data acquisition

Clinical data were obtained prospectively via clinical interaction with enrolled subjects at time of preoperative evaluation and postoperative clinical follow-up 3 months postoperatively by clinical investigators, based on clinical assessment of shunt surgery response in each of the three symptomatic domains of the NPH triad. All patients were seen in a NPH specialty clinic by attending neurosurgeons specialized in NPH and other CSF disorders (P.K. and K.S.). Data were entered into the electronic medical record as part of standard clinical documentation and later abstracted by study team members (O.P.L. and S.B.) for use as study endpoints.

### CSF sample processing

Patient CSF samples were collected intraoperatively during ventriculoperitoneal shunt placement procedures. Each 15 mL sample was procured immediately upon placement of the ventricular catheter into a sterile, chilled polypropylene collection tube. The sample was then immediately transferred on ice for processing and storage. 10 mL of the sample was centrifuged at 1500 relative centrifugal force (RCF) for 10 minutes at 4°C to remove debris. Supernatant was collected (∼10 mL), flash frozen in liquid nitrogen, and stored at −80°C for extracellular vesicle (EV) isolation and mRNA extraction. The remaining 5 mL of the sample was aliquoted into multiple 0.15 mL and 2 mL polypropylene storage tubes for immunoassay analysis. These samples did not undergo centrifugation and were stored at −80°C.

### ELISA analysis and literature comparison

CSF samples from 34 NPH patients were tested for quantitative determination of core neurodegeneration markers at Quanterix, (Billerica, MA, USA), using Single Molecule Array (Simoa) technology^45, 46^. All CSF samples were diluted based on internal standards and measured in duplicate. The mean of replicates for each sample was calculated and represented graphically. To compare levels of core neurodegeneration markers in this NPH cohort with published data from non-dementia controls and AD patients, we searched for literature reporting levels of neurodegeneration markers in CSF. Concentrations of Aβ42 in control patients were identified in^47^, and concentrations of Aβ42 in AD and NPH patients were identified in^28^. Concentrations of p-tau231 for control and AD patients were identified in^48^. Concentrations of IL-6 for control and AD patients in were identified in^49^. Concentrations of IL-8 in control and AD patients were identified in^50^. Concentrations of TNFα for control and AD patients were identified in^51^. Concentrations of GFAP for control patients (individuals with idiopathic intracranial hypertension) were identified in^52^, and concentrations for patients with AD were identified in^53^. Concentrations of NfL for control and AD patients were identified in^54^. Concentrations of IL-8 in NPH patients were identified in^55^.

### Extracellular vesicle (EV) isolation and mRNA extraction

EV isolation and mRNA extraction was performed at the Extracellular Vesicle Core Facility at Rhode Island Hospital. CSF was centrifuged at 2000 g for 20 minutes at room temperature. The samples were aliquoted and stored at −80°C. The 10 mL frozen CSF aliquots were thawed on ice and diluted 1:1 in PBS. EVs were isolated by ultracentrifugation in UltraClear 30 mL tubes at 100,000 g for 70 minutes at 4°C. They were resuspended in 100 µL of PBS supplemented with 1% DMSO and stored at −80°C. RNA was extracted using the miRNeasy Micro Kit according to the manufacturer’s protocol (Qiagen, ID: 217084).

### RNA sequencing and data analysis

cDNA library preparation and paired-end Illumina sequencing on a NovaSeq 6000 sequencing system were performed at The University of Chicago Genomics Facility. Samples were sequenced in two batches. The first set of samples was sequenced with two technical replicates per sample and the second set of samples was sequenced with three technical replicates per sample. FASTQ files were transferred to Brown University’s high-performance computing cluster OSCAR using the Globus file transfer system. “Trim Galore!” (version 0.5.0) was used to remove Illumina adapter sequences from the raw FASTQ files and FastQC (version 0.11.5) was used to assess the quality of the data. Sequencing reads were then aligned to the human transcriptome (Ensembl GrCh38 release 104) using HiSat2 (version 2.1.0) with the --dta flag^56^. Samtools (version 1.9) were used to convert and sort the sam files to bam files in preparation for mapping and alignment. StringTie (version 1.3.4) was used to assemble alignments into transcripts according to the differential expression workflow described in the StringTie manual^57, 58^. Each bam file was assembled into transcripts, the assembled transcripts were merged into a non-redundant set of transcripts across all samples, and the bam files were assembled against the merged transcript set to estimate transcript abundances and generate read coverage tables.

### Differential gene expression analysis

The estimated transcript abundances calculated by StringTie were imported into R (version 4.1.2) using the tximport package (version 1.22.0)^59^. Transcripts were summarized to the gene level using the tx2gene feature of the tximport function. Principal component analysis (PCA) of the top 1000 genes by mean expression was used to check for batch effects and detect outliers. Since technical replicates clustered together in PCA space, they were combined for all downstream analysis. The number of genes expressed in each sample was 8730.1 ± 2544.2. DESeq2 (version 1.34.0) was used for differential gene expression analysis^60^. All RNA samples were organized into a single DESeqDataSet using the DESeqDataSetFromTximport function. The design formula for each analysis included a sequencing batch variable plus the clinical outcome of interest. Genes that did not have more than 10 counts in at least 5 samples were filtered out before differential gene expression analysis. Differential gene expression analysis then followed the standard workflow for DESeq2.

### Gene set enrichment analysis

Gene set enrichment analysis was carried out using the GSEA tool (version 4.2.3)^61^. Genes were ranked using the Wald statistic calculated by DESeq2 and analyzed using the GSEA preRanked function. Kyoto Encyclopedia of Genes and Genomes (KEGG) pathways in the Molecular Signatures Database (MSigDB, version 7.5.1) were used. Pathway enrichment results were visualized and plotted using Python (version 3.10.2) and matplotlib (version 3.5.1). Pathways were considered enriched if they showed a FDR < 0.25.

### Feature selection and machine learning

Each 3-month clinical response was treated as an independent, binary classification task: gait improvement, urinary improvement, and cognitive improvement. Technical replicates were combined and samples missing the clinical response were excluded. RNA-seq counts transformed by the variance stabilizing transformation (vst) function in DESeq2 were exported from R into Python. The Python package scikit-learn (version 1.0.2) was used for all pre-processing and machine learning steps^62^. Leave-One-Out cross validation was used to generate N training and testing splits where N was the total number of samples. In each fold, gene expression was z-scored using the StandardScaler function in scikit-learn and genes were selected based on their weight in a logistic regression model. To test how the number of selected genes impacted classifier performance, we varied the number of selected genes between 10 and 1000. For each number of selected genes, a support vector machine (SVM) classifier with C = 1 was trained and evaluated. We used these model hyperparameters (SVM, C = 1) for all predictions as they showed good performance across the three clinical responses and our sample size prevented us from optimizing hyperparameters through cross-validation. To evaluate the classifier’s performance, each sample was held out as a test sample once and the predictions for all samples were concatenated before calculating the area under the receiver operating characteristic curve (AUC). A SVM trained on the smallest number of genes that produced the highest AUC was used for plotting a receiver operating characteristic (ROC) curve. PCA fit to the expression of the selected genes in the first training set was used to visualize the samples in two dimensions of the feature space.

### Statistical analyses

All statistical analysis was performed using R (version 4.1.2), RStudio (version 2022.02.0-443), and Python (version 3.10.2). Chi-square tests and two-sided Mann-Whitney U-tests with a significance threshold of p < 0.05 were used to test for differences in sex and age demographics (Supplementary Figure 1). For ELISA data in Figure 2, groups were compared using two-sided Mann-Whitney U-tests with a significance threshold of Benjamini-Hochberg adjusted p < 0.05. RNA-seq data in Figure 3 and 4 were analyzed in DESeq2 with a significance level of Benjamini-Hochberg adjusted p < 0.05. KEGG pathways were tested in GSEA and considered significantly enriched with a FDR < .25. Normalized expression of top differentially expressed genes was compared with a two-sided Mann-Whitney U-test with a significance threshold of p < 0.05.

## Data Availability

Anonymized study data including RNA sequencing data are available upon request. The R and Python analysis scripts used for this study are available at the GitLab link https://gitlab.com/nph_biomarkers.

## Acknowledgments

We thank Teah Markstone for help with RNA sequencing analysis, Sicheng Wen and the Lifespan COBRE Center for Stem Cells and Aging for the preparation of extracellular vesicles, and the University of Chicago Genomics Facility for RNA sequencing. We thank Rick Gerkin and Jason Machan for advice on statistics and Mark Albers and Justin Fallon for critical comments on the manuscript. This work was supported by a Zimmerman Innovation Award from the Carney Institute for Brain Science and a Brown University OVPR Research Seed Award.

## Author contributions

O.P.L., T.S., U.A., P.K., A.F. and M.G.R. conceived and designed this study; K.S. and P.K. performed shunt surgeries; O.L., S.B., K.S. and M.G.R. collected data; Z.L., V.M., S.K., T.S., and A.F. performed analyses; Z.L. and A.F. wrote the manuscript, with contributions from all authors.

## Competing interests

T. S., P.K. and A.F. are advisors and co-founders of Adelle Diagnostics, Inc. M.G.R. is the CEO and co-founder of Adelle Diagnostics, Inc. The remaining authors declare no competing interests.

---

**Supplementary Figure 1.**
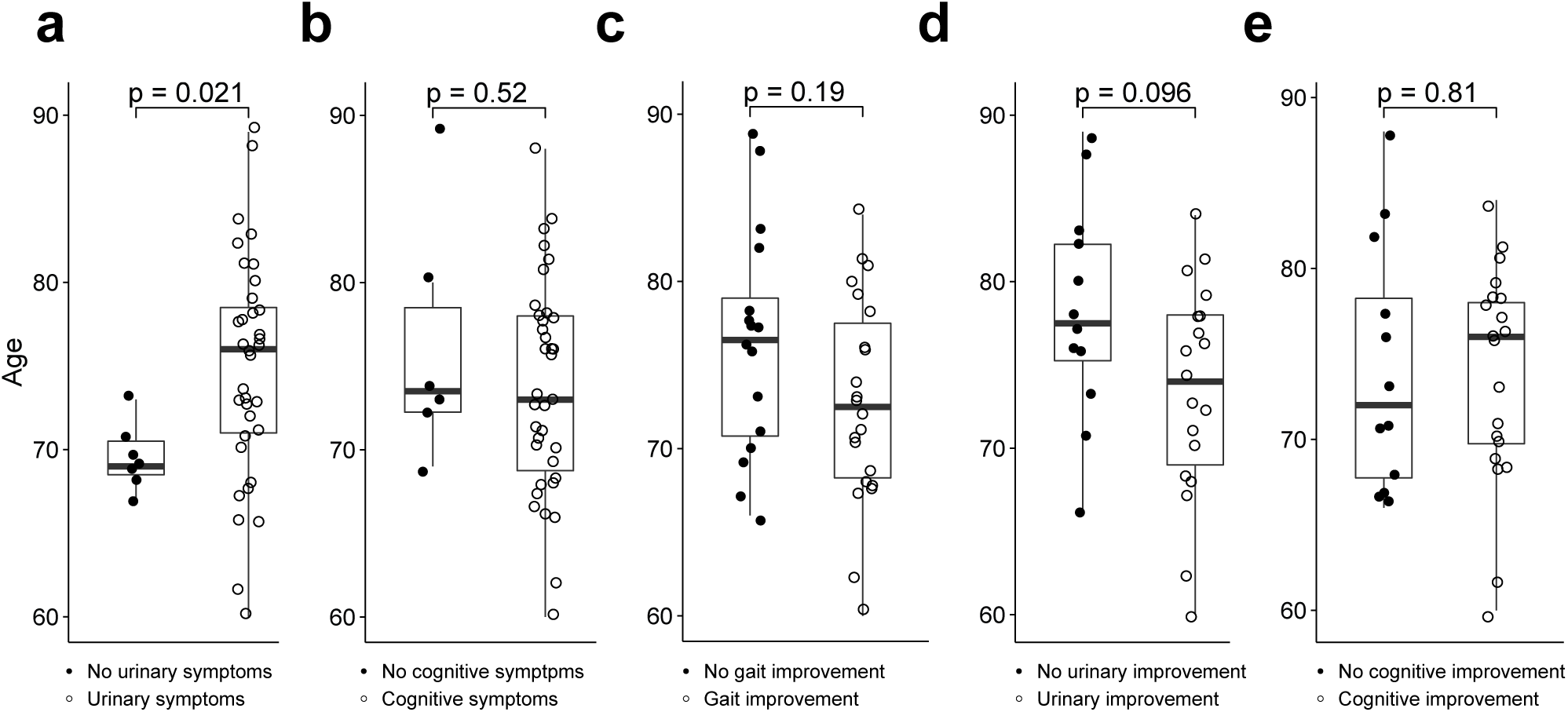
Patients with urinary symptoms, but not cognitive symptoms or postoperative gait, urinary, or cognitive improvement, show differences in age. **a.** Ages of patients with (n = 35) or without (n = 7) urinary symptoms before surgery. All U- and p-values calculated by two-sided Mann-Whitney U-test. U = 191, p = 0.02. **b.** Ages of patients with (n = 36) or without (n = 6) cognitive symptoms before surgery. U = 89.5, p = 0.52. **c.** Ages of patients with (n = 22) or without (n = 16) gait improvement after surgery. U = 131.5, p = 0.19. **d.** Ages of patients with (n =19) or without (n = 12) urinary symptom improvement after surgery. U = 72.5, p = 0.096. **e.** Ages of patients with (n = 20) or without (n = 12) cognitive improvement after surgery. U = 126.5, p = 0.81.

**Supplementary Figure 2:**
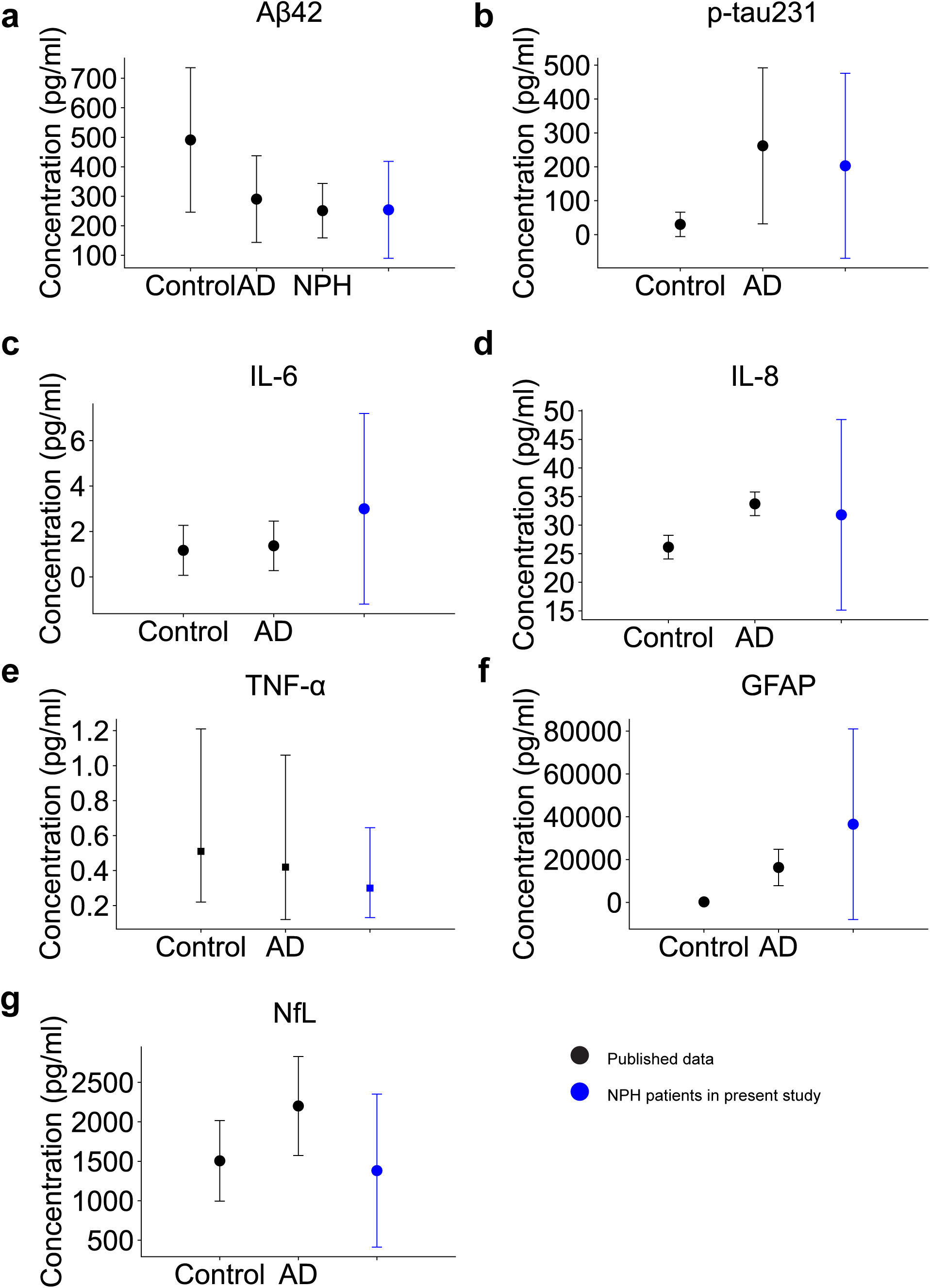
Comparisons between levels of core CSF biomarkers for neurodegeneration in this NPH cohort and published data for NPH, Alzheimer’s Disease, and non-dementia controls. **a.** Concentrations of Aβ42 in this NPH patient cohort (n = 32) compared to published values for healthy controls (n = 72), Alzheimer’s Disease (AD) patients (n = 11), and NPH patients (n =11)^28, 47^. Plotted as mean ± 1 standard deviation. Blue data: this NPH patient cohort. Black: published data. All published data are from CSF collected by lumbar puncture. Only mean and standard deviations of published data were available. To avoid assuming published data follow normal distributions, we did not perform statistical comparisons between groups. **b.** Concentrations of pTau231 in this NPH cohort (n = 25) compared to published values for healthy controls (n = 26) and AD patients (n = 21)^48^. Plotted as mean ± 1 standard deviation. **c.** Concentrations of IL-6 in this NPH cohort (n = 42) compared to published values for healthy controls (n = 36) and AD patients (n = 45)^49^. Plotted as mean ± 1 standard deviation. **d.** Concentrations of IL-8 in this NPH cohort (n = 42) compared to published values for healthy controls (n = 41) and AD patients (n = 36)^50, 55^. Plotted as mean ± 1 standard deviation. **e.** Concentrations of TNFα in this NPH cohort (n = 26) compared to published values for healthy controls (n = 23) and AD patients (n = 41)^51^. Data plotted as median and interquartile range. **f.** Concentrations of GFAP in this NPH cohort (n = 32) compared to published values for non-dementia controls (n = 39) and AD patients (n = 300)^52, 53^. Plotted as mean ± 1 standard deviation. **g.** Concentrations of NfL in this NPH cohort (n = 42) compared to published values for healthy controls (n = 159) and AD patients (n = 28)^54^. Plotted as mean ± 1 standard deviation.

**Supplementary Table 1:** Postoperative shunt responses for patients split by combinations of NPH symptoms reported preoperatively.

| **Combinations of Preoperative Symptoms Reported** | **Number (%)** | **Gait Symptoms Number, (%)** | **Urinary Symptoms Number, (%)** | **Cognitive Symptoms Number, (%)** | **Response After Shunt Surgery** |
| --- | --- | --- | --- | --- | --- |
| Gait & Urinary & Cognitive | 31 (73.8%) | 16 (51.6%) | 17 (54.8%) | 16 (51.6%) | Improved |
|  |  | 11 (35.5%) | 10 (32.2%) | 11 (35.5%) | Not Improved |
|  |  | 4 (12.9%) | 4 (12.9%) | 4 (12.9%) | Missing |
| Gait & Cognitive | 5 (11.9%) | 3 (60%) | – | 4 (80%) | Improved |
|  |  | 2 (40%) | – | 1 (20%) | Not Improved |
|  |  | 0 | – | 0 | Missing |
| Gait & Urinary | 4 (9.5%) | 3 (75%) | 2 (50%) | – | Improved |
|  |  | 1 (25%) | 2 (50%) | – | Not Improved |
|  |  | 0 | 0 | – | Missing |
| Gait Only | 2 (4.8%) | 0 | – | – | Improved |
|  |  | 2 (100%) | – | – | Not Improved |
|  |  | 0 | – | – | Missing |

**Supplementary Table 2:** Differentially expressed genes for urinary symptoms, cognitive symptoms, and postoperative cognitive improvement.

| Clinical Trait | Differentially Expressed Genes |
| --- | --- |
| Preoperative urinary symptoms | *MOK, NID1, BLVRB, RMND5B, KLHL11, TKFC, SPEF1, CYP26B1, CLEC9A, BCKDHA, TPPP, ARMCX3, IGFBP6, IGFN1, KIAA1549L, LINC00911, ZNF471, GCLM, MYH3, RIT1, TTC30B, NNT, KLHL28, LIN28A, CMTR2, SPRY1, TMEM70, ZIC4, OTULINL, TENT4B, MTX3, NRSN2, IQGAP2, LIG4, TRMT5, CCNL1, NFKBIA, NIPSNAP3A, SERPIND1, SIX4, SCN1A, OMD, PIH1D2, USP5, CDADC1, PPP1R12C, FSTL1, ZCCHC8, SLC6A20, HOMEZ, PGS1, REL, KLHL32, S100A4, ANKRD36BP2, PASK, GABBR2, EML2, ALDH9A1, POLR2A, RING1, ZXDA, SHC3, FAM104A, C16orf87, GORAB, IAH1, OGFR* |
| Preoperative cognitive symptoms | *HBG1, ST3GAL3, USP5* |
| Postoperative cognitive improvement | *TP53I11, TRIM52-AS1, BIRC3, PIM1, ALC22A15, ANAPC10, TFEC* |

**Supplementary Table 3.** Candidate RNA biomarkers based on consistent selection by machine learning classifier. Lists of genes selected by weight in the logistic regression model in all folds of the classifiers predicting gait, urinary, and cognitive improvement.

| Improvement 3-Months<br>After Shunting | Genes Selected In Every Classifier Fold<br>(constant/total per fold, % of each fold) |
| --- | --- |
| Gait | *AP4E1, CLIC2, COL6A2, FAM160B2, HCP5,*<br>*KCTD7, LRRK2, PPP1R3E, PRKAB1, RGS12,*<br>*RTN4IP1, SAXO2*<br>(12/30, 40%) |
| Urinary | *ABLIM3, ACADM, ASAP3, BTN2A2, CCDC173,*<br>*CD22, CERCAM, CHIC2, EMC2, ERLIN1,*<br>*FAM47E, STBD1, GAREM1, GATAD2A, GLIPR1,*<br>*GSN-AS1, HINT2, HPS6, HYLS1, KCTD16,*<br>*KLHL42, LGMN, LINC00472, MAP3K21,*<br>*MFSD14A, MORN1, MRPL21, MVD, NNT-AS1,*<br>*NSUN4, OAS2, OVCH1-AS1, PHKG1, RN7SKP71,*<br>*SAXO2, SLC20A1, SMN1, TAX1BP3, TOB1-AS1,*<br>*TRAF1, WSCD2, ZDHHC1, ZNF708*<br>(43/140, 31%) |
| Cognition | *BEND3P3, CERS6, ENOX2, JCAD, KPNA4,*<br>*PKNOX1, POLR2J4, RNU4ATAC, SCRT1,*<br>*SH3TC2, TMEM202-AS1, UTP20*<br>(12/50, 24%) |

## Notes

### Funding Statement

This work was supported by a Zimmerman Innovation Award from the Carney Institute for Brain Science at Brown University, and a Brown University OVPR Research Seed Award.

### Author Declarations

The Institutional Review Board (IRB) of the Rhode Island Hospital gave ethical approval for this work

## References

1. Dugger, B. N. & Dickson, D. W. Pathology of Neurodegenerative Diseases. Cold Spring Harb. Perspect. Biol. 9, a028035 (2017).

2. Espay, A. J. et al. Deconstructing normal pressure hydrocephalus: Ventriculomegaly as early sign of neurodegeneration. Ann. Neurol. 82, 503–513 (2017).

3. Agrawal, M. & Biswas, A. Molecular diagnostics of neurodegenerative disorders. Front. Mol. Biosci. 2, 54 (2015).

4. Doroszkiewicz, J., Groblewska, M. & Mroczko, B. Molecular Biomarkers and Their Implications for the Early Diagnosis of Selected Neurodegenerative Diseases. Int. J. Mol. Sci. 23, 4610 (2022).

5. Siraj, S. An overview of normal pressure hydrocephalus and its importance: how much do we really know? J. Am. Med. Dir. Assoc. 12, 19–21 (2011).

6. Martín-Láez, R., Caballero-Arzapalo, H., López-Menéndez, L. Á., Arango-Lasprilla, J. C. & Vázquez-Barquero, A. Epidemiology of Idiopathic Normal Pressure Hydrocephalus: A Systematic Review of the Literature. World Neurosurg. 84, 2002–2009 (2015).

7. Andersson, J. et al. Prevalence of idiopathic normal pressure hydrocephalus: A prospective, population-based study. PloS One 14, e0217705 (2019).

8. Kiefer, M. & Unterberg, A. The differential diagnosis and treatment of normal-pressure hydrocephalus. Dtsch. Arzteblatt Int. 109, 15–25; quiz 26 (2012).

9. Tseng, P.-H. et al. Diagnosis and treatment for normal pressure hydrocephalus: From biomarkers identification to outcome improvement with combination therapy. Tzu Chi Med. J. 34, 35–43 (2022).

10. Jaraj, D. et al. Prevalence of idiopathic normal-pressure hydrocephalus. Neurology 82, 1449–1454 (2014).

11. Hebb, A. O. & Cusimano, M. D. Idiopathic normal pressure hydrocephalus: a systematic review of diagnosis and outcome. Neurosurgery 49, 1166–1184; discussion 1184-1186 (2001).

12. Shprecher, D., Schwalb, J. & Kurlan, R. Normal pressure hydrocephalus: diagnosis and treatment. Curr. Neurol. Neurosci. Rep. 8, 371–376 (2008).

13. Klinge, P., Marmarou, A., Bergsneider, M., Relkin, N. & Black, P. M. Outcome of shunting in idiopathic normal-pressure hydrocephalus and the value of outcome assessment in shunted patients. Neurosurgery 57, S40–52; discussion ii-v (2005).

14. Miyajima, M., Kazui, H., Mori, E., Ishikawa, M., & , on behalf of the SINPHONI-2 Investigators. One-year outcome in patients with idiopathic normal-pressure hydrocephalus: comparison of lumboperitoneal shunt to ventriculoperitoneal shunt. J. Neurosurg. 125, 1483–1492 (2016).

15. Vakili, S. et al. Timing of surgical treatment for idiopathic normal pressure hydrocephalus: association between treatment delay and reduced short-term benefit. Neurosurg. Focus 41, E2 (2016).

16. Feletti, A. et al. Ventriculoperitoneal Shunt Complications in the European Idiopathic Normal Pressure Hydrocephalus Multicenter Study. Oper. Neurosurg. Hagerstown Md 17, 97–102 (2019).

17. Johanson, C. E. et al. Multiplicity of cerebrospinal fluid functions: New challenges in health and disease. Cerebrospinal Fluid Res. 5, 10 (2008).

18. Spector, R., Keep, R. F., Robert Snodgrass, S., Smith, Q. R. & Johanson, C. E. A balanced view of choroid plexus structure and function: Focus on adult humans. Exp. Neurol. 267, 78–86 (2015).

19. Telano, L. N. & Baker, S. Physiology, Cerebral Spinal Fluid. in StatPearls (StatPearls Publishing, 2022).

20. Tumani, H. et al. Cerebrospinal fluid biomarkers of neurodegeneration in chronic neurological diseases. Expert Rev. Mol. Diagn. 8, 479–494 (2008).

21. Saugstad, J. A. et al. Analysis of extracellular RNA in cerebrospinal fluid. J. Extracell. Vesicles 6, 1317577 (2017).

22. Darrow, J. A. et al. CSF Biomarkers Predict Gait Outcomes in Idiopathic Normal Pressure Hydrocephalus. Neurol. Clin. Pract. 12, 91–101 (2022).

23. Jeppsson, A. et al. CSF biomarkers distinguish idiopathic normal pressure hydrocephalus from its mimics. J. Neurol. Neurosurg. Psychiatry 90, 1117–1123 (2019).

24. Taghdiri, F. et al. Association Between Cerebrospinal Fluid Biomarkers and Age-related Brain Changes in Patients with Normal Pressure Hydrocephalus. Sci. Rep. 10, 9106 (2020).

25. Pfanner, T., Henri-Bhargava, A. & Borchert, S. Cerebrospinal Fluid Biomarkers as Predictors of Shunt Response in Idiopathic Normal Pressure Hydrocephalus: A Systematic Review. Can. J. Neurol. Sci. J. Can. Sci. Neurol. 45, 3–10 (2018).

26. Manniche, C. et al. Cerebrospinal Fluid Biomarkers to Differentiate Idiopathic Normal Pressure Hydrocephalus from Subcortical Ischemic Vascular Disease. J. Alzheimers Dis. JAD 75, 937–947 (2020).

27. Blum-Degen, D. et al. Interleukin-1 beta and interleukin-6 are elevated in the cerebrospinal fluid of Alzheimer’s and de novo Parkinson’s disease patients. Neurosci. Lett. 202, 17–20 (1995).

28. Michael MalekAhmadi, A. T. Differences in Cerebrospinal Fluid Biomarkers between Clinically Diagnosed Idiopathic Normal Pressure Hydrocephalus and Alzheimer’s Disease. J. Alzheimers Dis. Park. 04, (2014).

29. Rydbirk, R. et al. Cytokine profiling in the prefrontal cortex of Parkinson’s Disease and Multiple System Atrophy patients. Neurobiol. Dis. 106, 269–278 (2017).

30. Chen, X., Hu, Y., Cao, Z., Liu, Q. & Cheng, Y. Cerebrospinal Fluid Inflammatory Cytokine Aberrations in Alzheimer’s Disease, Parkinson’s Disease and Amyotrophic Lateral Sclerosis: A Systematic Review and Meta-Analysis. Front. Immunol. 9, 2122 (2018).

31. Hu, W. T. et al. CSF Cytokines in Aging, Multiple Sclerosis, and Dementia. Front. Immunol. 10, 480 (2019).

32. Tullberg, M. et al. Cerebrospinal fluid markers before and after shunting in patients with secondary and idiopathic normal pressure hydrocephalus. Cerebrospinal Fluid Res. 5, 9 (2008).

33. Ishiki, A. et al. Glial fibrillar acidic protein in the cerebrospinal fluid of Alzheimer’s disease, dementia with Lewy bodies, and frontotemporal lobar degeneration. J. Neurochem. 136, 258–261 (2016).

34. Bartl, M. et al. Biomarkers of neurodegeneration and glial activation validated in Alzheimer’s disease assessed in longitudinal cerebrospinal fluid samples of Parkinson’s disease. PloS One 16, e0257372 (2021).

35. Schulz, I. et al. Systematic Assessment of 10 Biomarker Candidates Focusing on α-Synuclein-Related Disorders. Mov. Disord. Off. J. Mov. Disord. Soc. 36, 2874–2887 (2021).

36. Hashimoto, M., Ishikawa, M., Mori, E., Kuwana, N., & Study of INPH on neurological improvement (SINPHONI). Diagnosis of idiopathic normal pressure hydrocephalus is supported by MRI-based scheme: a prospective cohort study. Cerebrospinal Fluid Res. 7, 18 (2010).

37. Noori, A., Mezlini, A. M., Hyman, B. T., Serrano-Pozo, A. & Das, S. Systematic review and meta-analysis of human transcriptomics reveals neuroinflammation, deficient energy metabolism, and proteostasis failure across neurodegeneration. Neurobiol. Dis. 149, 105225 (2021).

38. Prince, M. J. et al. World Alzheimer Report 2015 - The Global Impact of Dementia: An analysis of prevalence, incidence, cost and trends. (2015).

39. Allali, G., Laidet, M., Armand, S. & Assal, F. Brain comorbidities in normal pressure hydrocephalus. Eur. J. Neurol. 25, 542–548 (2018).

40. Purro, S. A., Galli, S. & Salinas, P. C. Dysfunction of Wnt signaling and synaptic disassembly in neurodegenerative diseases. J. Mol. Cell Biol. 6, 75–80 (2014).

41. Singh, A., Kukreti, R., Saso, L. & Kukreti, S. Oxidative Stress: A Key Modulator in Neurodegenerative Diseases. Mol. Basel Switz. 24, E1583 (2019).

42. Schmidt, M. F., Gan, Z. Y., Komander, D. & Dewson, G. Ubiquitin signalling in neurodegeneration: mechanisms and therapeutic opportunities. Cell Death Differ. 28, 570– 590 (2021).

43. Sato, H. et al. A Segmental Copy Number Loss of the SFMBT1 Gene Is a Genetic Risk for Shunt-Responsive, Idiopathic Normal Pressure Hydrocephalus (iNPH): A Case-Control Study. PloS One 11, e0166615 (2016).

44. Yang, H. W. et al. Deletions in *CWH43* cause idiopathic normal pressure hydrocephalus. EMBO Mol. Med. 13, (2021).

45. Rissin, D. M. et al. Single-molecule enzyme-linked immunosorbent assay detects serum proteins at subfemtomolar concentrations. Nat. Biotechnol. 28, 595–599 (2010).

46. Wilson, D. H. et al. The Simoa HD-1 Analyzer: A Novel Fully Automated Digital Immunoassay Analyzer with Single-Molecule Sensitivity and Multiplexing. J. Lab. Autom. 21, 533–547 (2016).

47. Sunderland, T. et al. Decreased beta-amyloid1-42 and increased tau levels in cerebrospinal fluid of patients with Alzheimer disease. JAMA 289, 2094–2103 (2003).

48. Pilotto, A. et al. Differences between plasma and CSF p-tau181 and p-tau231 in early Alzheimer’s disease. http://medrxiv.org/lookup/doi/10.1101/2021.12.10.21267467 (2021) doi:10.1101/2021.12.10.21267467.

49. Wennström, M. et al. Cerebrospinal fluid levels of IL-6 are decreased and correlate with cognitive status in DLB patients. Alzheimers Res. Ther. 7, 63 (2015).

50. Galimberti, D. et al. Intrathecal Chemokine Synthesis in Mild Cognitive Impairment and Alzheimer Disease. Arch. Neurol. 63, 538 (2006).

51. Hesse, R. et al. Decreased IL-8 levels in CSF and serum of AD patients and negative correlation of MMSE and IL-1β. BMC Neurol. 16, 185 (2016).

52. Michel, M. et al. Increased GFAP concentrations in the cerebrospinal fluid of patients with unipolar depression. Transl. Psychiatry 11, 308 (2021).

53. Benedet, A. L. et al. Differences Between Plasma and Cerebrospinal Fluid Glial Fibrillary Acidic Protein Levels Across the Alzheimer Disease Continuum. JAMA Neurol. 78, 1471 (2021).

54. Dhiman, K. et al. Cerebrospinal fluid neurofilament light concentration predicts brain atrophy and cognition in Alzheimer’s disease. Alzheimers Dement. Diagn. Assess. Dis. Monit. 12, (2020).

55. Pyykkö, O. T. et al. Cerebrospinal fluid biomarker and brain biopsy findings in idiopathic normal pressure hydrocephalus. PloS One 9, e91974 (2014).

56. Kim, D., Paggi, J. M., Park, C., Bennett, C. & Salzberg, S. L. Graph-based genome alignment and genotyping with HISAT2 and HISAT-genotype. Nat. Biotechnol. 37, 907–915 (2019).

57. Pertea, M. et al. StringTie enables improved reconstruction of a transcriptome from RNA-seq reads. Nat. Biotechnol. 33, 290–295 (2015).

58. Pertea, M., Kim, D., Pertea, G. M., Leek, J. T. & Salzberg, S. L. Transcript-level expression analysis of RNA-seq experiments with HISAT, StringTie and Ballgown. Nat. Protoc. 11, 1650–1667 (2016).

59. Soneson, C., Love, M. I. & Robinson, M. D. Differential analyses for RNA-seq: transcript-level estimates improve gene-level inferences. Preprint at https://doi.org/10.12688/f1000research.7563.2 (2016).

60. Love, M. I., Huber, W. & Anders, S. Moderated estimation of fold change and dispersion for RNA-seq data with DESeq2. Genome Biol. 15, 550 (2014).

61. Subramanian, A. et al. Gene set enrichment analysis: A knowledge-based approach for interpreting genome-wide expression profiles. Proc. Natl. Acad. Sci. 102, 15545–15550 (2005).

62. Pedregosa, F. et al. Scikit-learn: Machine Learning in Python. J. Mach. Learn. Res. 12, 2825–2830 (2011).

